# Immunophenotyping of B- and T-cell alterations in patients with autoimmune premature ovarian insufficiency following rituximab treatment

**DOI:** 10.64898/2026.09.22.26363548

**Authors:** Kittikorn Wangriatisak, Annelien Hooijsma, Kajsa Amnehagen, Francesca Faustini, Maribel Aranda-Guillén, Sigridur Björnsdottir, Sophie Bensing, Olle Kämpe, Angelica Lindén Hirschberg, Iva Gunnarsson, Vivianne Malmström

**Affiliations:** Division of Rheumatology, Department of Medicine Solna, Karolinska Institutet, Karolinska University Hospital, Sweden; Center for Molecular Medicine, Karolinska Institutet, Sweden; Department of Endocrinology, Karolinska Institutet, Karolinska University Hospital, Sweden; Department of Medicine, Solna, Karolinska Institutet, Sweden; Department of Women’s and Children’s Health, Karolinska Institutet, Sweden; Department of Gynecology and Reproductive Medicine, Karolinska University Hospital, Sweden

**Author notes:** Corresponding author details: Vivianne Malmström, PhD, Division of Rheumatology, Department of Medicine, Karolinska Institutet, Karolinska University Hospital Solna, 171 77, Stockholm, Sweden.

## Abstract

**Background:** Premature ovarian insufficiency (POI) is a condition affecting female fertility e.g., in the context of autoimmune disease. A recent proof-of-concept study showed that B-cell depletion with rituximab (RTX) temporarily restored ovarian function in patients with autoimmune POI. However, the underlying immunological mechanisms behind this response is unknown.

**Objectives:** To characterize B- and T-cell phenotypes before and after RTX therapy in the above-mentioned patients and associations with autoantibody levels and treatment response.

**Methods:** Cryopreserved cell samples were available from six autoimmune patients with POI, four of whom were positive for autoantibodies against steroidogenic enzymes (21OH, 17αOH and/or SCC). Patients received RTX and blood samples were collected at baseline, second infusion (2 weeks), 3, 8 and 12 months. Peripheral B- and T-cell phenotypes were analysed using spectral flow cytometry and correlated with autoantibody levels.

**Results:** At baseline, patients displayed signs of B-cell activation by alteration in subset composition compared to matched controls. Upon closer analysis, patients positive for steroidogenic enzyme autoantibodies had significantly increased frequency of CXCR3+switched memory (SWM) B-cells than autoantibody-negative patients, which correlated positively with anti-17αOH and-SCC autoantibody levels. In parallel, increased Th1 cell frequencies were also observed in the same patient group. Longitudinal analyses showed expected B-cell depletion followed by repopulation after RTX. Changes in T-cell subsets were observed at 8 months with reduced frequencies of Th1 and Tfh cells.

**Conclusion:** Autoimmune POI displayed altered B- and T-cell subsets implicating cell crosstalk, immune cell infiltration to sites of inflammation such as ovaries, and presence of Th1-driven immunopathogenesis.

*Key messages:*

- Increased CXCR3+SWM B cells and Th1 cells were found in patients with autoimmune POI who were positive for autoantibodies against steroidogenic enzymes.
- CXCR3 expression on SWM B cells positively associated with anti-17αOH and anti-SCC autoantibody levels.
- RTX induced rapid B-cell depletion followed by repopulation and a delayed T-cell effect at 8 months with reduced Th1 and Tfh cell frequencies.

*Capsule summary:* RTX has recently been shown to restore fertility in autoimmune POI. Lymphocyte profiling identified Th1 bias accompanied by CXCR3+memory B cells that correlate with autoantibody levels. B-cell depletion appears to temporarily alleviate ovarian autoimmunity and ameliorate physiological function.

## Introduction

Premature ovarian insufficiency (POI) is defined as the loss of ovarian function before the age of 40 and is a major cause of female infertility, affecting approximately 3.5% of women worldwide (1). The aetiology of POI is heterogeneous with genetic, infectious and autoimmune factors. Autoimmunity is estimated to account for approximately 5-30% of all POI cases (2,3), with autoimmune POI being frequently associated with other autoimmune diseases e.g., autoimmune adrenal insufficiency and autoimmune polyglandular syndromes (3,4). Autoantibodies against steroidogenic enzymes such as 21-hydroxylase (21OH), 17α-hydroxylase (17αOH) and cholesterol side-chain cleavage enzyme (SCC) and in some cases thyroperoxidase have been detected in patients with autoimmune POI (5–9). The presence of these autoantibodies implicates dysregulated humoral immune responses.

B-cell targeted therapies have demonstrated efficacy in several autoimmune diseases e.g., systemic lupus erythematosus (SLE), rheumatoid arthritis (RA) and multiple sclerosis (MS) (10–12). In autoimmune POI, a recent proof-of-concept study demonstrated successful oocyte retrieval in response to ovarian stimulation and subsequent pregnancies in patients treated with rituximab (RTX), an anti-CD20 monoclonal antibody (13). In addition to B-cell depletion, RTX has also been shown to modulate T-cell responses (14,15).

B cells comprise of functionally distinct subsets that undergo differentiation from transitional and naïve B cells into memory B cells and plasmablasts/cells (16–18). In addition to antibody production, B cells also function as antigen-presenting cells for autoreactive CD4+T cells and secrete inflammatory cytokines that exacerbate local inflammation (19, 20). Many systemic autoimmune diseases display various degrees of aberrations in distinct B-cell subpopulations that are associated with disease activity and/or autoantibody production such as double negative 2 (DN2) B cells which are abundant e.g., in SLE and RA (21–24). To our knowledge, such atypical B cells or DN2 cells have not yet been studied in autoimmune POI or in Addison’s disease which is the primary diagnosis for many individuals with autoimmune POI.

Previous studies in heterogeneous POI cohorts have implicated a T-cell imbalance characterized by deficiency of regulatory T (Tregs) and T helper 22 (Th22) cells together with augmented T helper 1 (Th1) cell responses (25–27). Despite accumulating evidence suggesting the involvement of both humoral and cellular immunity in autoimmune POI, the composition of B-cell compartments, their relationship with T-cell responses and contribution to disease development remain underexplored.

In this pilot study, we aimed to characterize circulating B- and T-cell compartments in patients with autoimmune POI using longitudinally cryopreserved peripheral blood mononuclear cells (PBMC) samples collected before and after RTX. We further explored their associations with autoantibody levels and immune changes upon RTX treatment.

## Methods

### Patients and samples

Patients with hypergonadotropic hypogonadism (FSH > 40 IU/L) and evidence of autoimmunity were recruited for a proof-of-concept study investigating whether RTX therapy could rescue ovarian function (13). Evidence of autoimmunity included autoantibodies against the 21OH, 17αOH and SCC (CYP11A1) or other autoantibody positive conditions. Participants were 29-34 years of age and had a body mass index (BMI) of 20-28 kg/m^2^. Patients were required to use non-hormonal contraception for 12 months following the final RTX treatment. Data collected included demographics, clinical manifestations, and laboratory parameters. Patients with chromosomal aberrations/abnormalities or fragile X premutation were excluded. Blood samples were available from six patients with autoimmune POI. All patients received RTX treatment and were sampled longitudinally at baseline, at the second infusion (2 weeks), and at 3, 8 and 12 months. Clinical outcomes following RTX treatment was available and was assessed based on recovery of ovarian function. Three patients were classified as responders while the remaining patients showed no or incomplete recovery of ovarian function. Samples were analysed for B cell and T cell immunophenotyping, followed by correlation analyses with serum autoantibody levels. Samples from age and sex-matched healthy control blood donors (HC, n = 6) were included as comparison.

### B cell phenotyping by spectral flow cytometry

PBMCs were isolated by Ficoll-Paque (Cytiva) density-gradient centrifugation and surface stained with fluorochrome-conjugated antibodies for 30 mins at 4°C (Table S1). After washing with 1xPBS, cells were co-stained with fixable viability dye eFluor 506 for 10 mins at 4°C. (1:1000, Invitrogen). Data were acquired using the Cytek Aurora spectral flow cytometer (5-laser, 355 nm, 405 nm, 488 nm, 561 nm and 640 nm). Both supervised and unsupervised analyses were performed. Live CD19+B cells were divided into switched memory (SWM: CD27+IgD-), unswitched memory (USW: CD27+IgD+), double negative (DN: CD27-IgD-), naïve (NAV: CD27-IgD+), transitional (TR: CD27-IgD+CD24hiCD38hi) and plasmablasts (PB: CD27hiCD38hi). Based on the expression of CD21 and CD11c, DN B cells were further subdivided into DN1 (CD11c-CD21+), DN2 (CD11c+CD21-) and DN3 (CD11c-CD21-), and NAV B cells into activated (CD11c+CD21-) and resting naive (CD11c-CD21+) (Figure S1).

### T cell phenotyping by spectral flow cytometry

PBMCs were surface stained with fluorochrome-conjugated antibodies and viability dye as described above (Table S2). After washing, cells were then fixed, permeabilised and intracellularly stained with fluorochrome-conjugated antibodies specific for CTLA-4 (BUV805), GZMK (AF647), GZMB (FITC) and FOXP3 (PE/Dazzle594) for 30 mins at room temperature (RT), and followed by acquisition using spectral flow cytometer. From Live CD3+T cells, cells were classified as TCR γδ+ and TCR γδ-. CD4+ and CD8+T cells were identified from the γδ-population. Both CD4+ and CD8+T cells were divided into central memory (CM: CCR7+CD45RA-), effector memory (EM: CCR7-CD45RA-), terminally differentiated effector memory (TEMRA: CCR7-CD45RA+) and naïve (NAV: CCR7+CD45RA+). CD4+T cells were further subdivided into T helper cell 1 (Th1: CCR6-CXCR3+CCR4-), Th2 (CCR6-CXCR3-CCR4+), Th17 (CCR6+CXCR3-), T peripheral helper cell (Tph-like: PD1hiCXCR3-HLA-DR+), T follicular helper cell (Tfh: PD1+CXCR5+), and regulatory T cell (Treg: FOXP3+) (Figure S2).

### Analyses of autoantibodies

Serum titers of autoantibodies against 21OH, 17αOH and SCC were measured in six women with autoimmune POI at baseline (pre-treatment), at the second RTX infusion and at 3, 8 and 12 months after treatment using a radioligand binding assay (28). Autoantibody levels were represented as relative index (RI) values.

## Statistical analysis

Data were analysed using GraphPad Prism 9.5.0 software and are represented as median (Q1-Q3). Medians were compared by Mann-Whitney U test (two groups) or Kruskal-Wallis (> 2 groups) with Dunn’s correction for multiple comparison test. Spearman’s test was applied to investigate correlations. For comparison of baseline and follow up samples, Wilcoxon-matched pairs signed rank test was used. P-values less than 0.05 were considered statistically significant.

## Ethics and consent

The study was approved by the Ethical Committee (2017/1398-31/1, 2018/1400-32, 2019/01825) and the Swedish Medical Products Agency (Dnr 5.1-2018-27443). Patients gave written informed consent to participate. The study was conducted in accordance with the Declaration of Helsinki.

## Results

### Patient characteristics

Patient characteristics are presented in Table 1. Four of six patients with autoimmune POI had autoimmune Addison’s disease. Of those, one patient also had type 1 diabetes, Hashimoto thyroiditis and celiac disease, while another one had alopecia, with multiple autoantibodies including 21OH, 17αOH and SCC autoantibodies. The two patients without Addison’s disease had Hashimoto thyroiditis and myasthenia gravis, respectively, with corresponding autoantibodies against thyroid peroxidase (TPO) and acetylcholine receptor (AChR). The mean age of patients at inclusion was 32.5 years and mean age at POI diagnosis was 26.2 years.

**Table 1.**

| Patient characteristics | Patients (n = 6) |
| --- | --- |
| Age at first autoimmune manifestation – yr | 22.5±5.5 |
| Gender, Female– n (%) | 6 (100) |
| Autoimmune manifestation |  |
| Addison’s disease – n (%) | 4 (66) |
| Hashimoto thyroiditis – n (%) | 2 (33) |
| Celiac disease – n (%) | 1 (16) |
| Alopecia – n (%) | 1 (16) |
| Myasthenia gravis – n (%) | 1 (16) |
| Type 1 diabetes – n (%) | 1 (16) |
| Autoantibodies |  |
| 21OH autoantibody positivity – n (%) | 3 (50) |
| SCC autoantibody positivity – n (%) | 4 (66) |
| 17αOH autoantibody positivity – n (%) | 3 (50) |
| TPO autoantibody positivity – n (%) | 2 (33) |
| AChR autoantibody positivity – n (%) | 1 (16) |
| Ages at POI diagnosis – yr | 26.2±4.9 |
| Ages at inclusion – yr | 32.5±1.8 |
| BMI at inclusion | 15.1±9.5 |
| Responder | 3 (50) |
| Pregnancy | 1 (17) |

Data are represented as mean and standard deviation or numbers and percentage.
Each patient may have multiple autoimmune manifestations
Each patient may have several autoantibodies
21-hydroxylase (21OH), cholesterol side-chain cleavage enzyme (SCC), 17α-hydroxylase (17αOH), thyroid peroxidase (TPO) and acetylcholine receptor (AChR), yr (year)
Responder: successful oocyte retrieval at ovarian stimulation

### Alteration of circulating B cell subsets in autoimmune POI patients

We first investigated the distribution of circulating B cell subsets in all autoimmune POI patients. The frequencies of total CD19+B cells and main subsets including SWM, USW, DN, NAV, TR and PB did not differ between patients with autoimmune POI and HC (Figure 1A, 1B and S3). Further analysis of DN and NAV subpopulations revealed increased frequencies of aNAV and DN3 in patients compared with HC (aNAV, *p* = 0.02 and DN3, *p* = 0.002), with DN2 also showed a similar trend. No differences were observed in DN1 and rNAV subsets (Figure 1B and S3).

**Figure 1.**
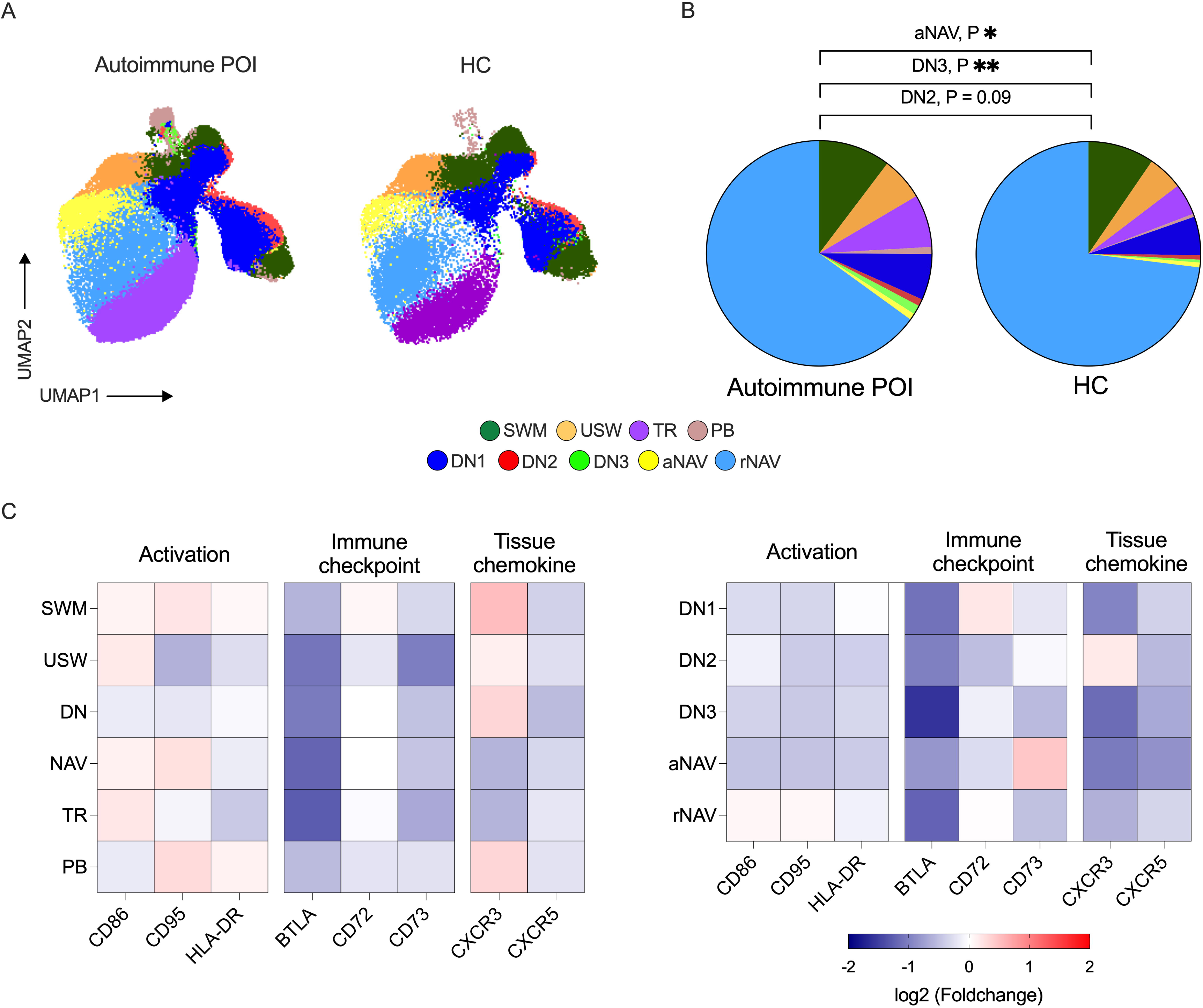
The distribution of B-cell subsets and their expression profiles in blood of patients with autoimmune POI. (A) Overlay of uniform manifold approximation and projection (UMAP) of clustering of CD19+B cells (n = 100,000 events each per patient and healthy control) and gated B cell subsets including switched memory (SWM: CD19+CD27+IgD-), unswitched memory (USW: CD19+CD27+IgD+), transitional (TR: CD19+CD27-IgD+CD24hiCD38hi), plasmablasts (PB: CD19+CD27+hiCD38hi), double negative 1 (DN1: CD19+CD27-IgD-CD11c-CD21+), double negative 2 (DN2: CD19+CD27-IgD-CD11c+CD21-), double negative 3 (DN3: CD19+CD27-IgD-CD11c-CD21-), activated naïve (aNAV: CD19+CD27-IgD+CD11c+CD21-) and resting naïve (rNAV: CD19+CD27-IgD+CD11c-CD21+). (B) Proportions of SWM, USW, TR, PB, DN1, DN2, DN3, aNAV and rNAV in patients with autoimmune POI (n = 6) compared to healthy controls (HC, n = 6). (C) Heatmap showing fold changes in the median fluorescence intensity (MFI) of activation (CD86, CD95 and HLA-DR), immune checkpoint (BTLA, CD72 and CD73) and tissue-chemokine (CXCR3 and CXCR5) expression on SWM, USW, DN, NAV, TR and PB (left panel) as well as DN1, DN2, DN3, aNAV and rNAV (right panel) from patients relative to HC. Data are represented as median and are analysed by Mann-Whitney U test. \**p*< 0.05; \*\**p*< 0.01; \*\*\**p*< 0.001; \*\*\*\**p*< 0.0001.

To further characterize B cell profiles, we assessed the expression of activation markers (CD86, CD95 and HLA-DR), immune checkpoint molecules (BTLA, CD72 and CD73) and tissue chemokine receptors (CXCR3 and CXCR5) in blood B cell subsets from autoimmune POI and HC (Figure 1C). Although none of the observed differences reached statistical significance, patients displayed numerically lower expression of immune checkpoint molecules especially BTLA and higher expression of CXCR3 in memory B cells (SWM, DN and DN2) and PB compared with HC.

### Reduced memory B cells and plasmablasts in patients positive for steroidogenic autoantibodies

We next investigated whether B cell subset distributions differed according to steroidogenic autoantibody status. Patients positive for at least one steroidogenic autoantibody (21OH, 17αOH and/or SCC; steroidogenic autoantibody-positive group) showed a marked reduction in SWM together with reduced plasmablast (PB) frequencies compared to steroidogenic autoantibody-negative patients. Compared with HC, steroidogenic autoantibody-negative patients showed a significant increase in DN3 B cell frequencies (*p* = 0.03), while SWM, USW, PB, NAV and aNAV cell frequencies also tended to be different, although these differences did not reach statistical significance. Patients positive for steroidogenic autoantibodies displayed increased frequencies of DN3 and aNAV B cells (Figure 2A).

**Figure 2.**
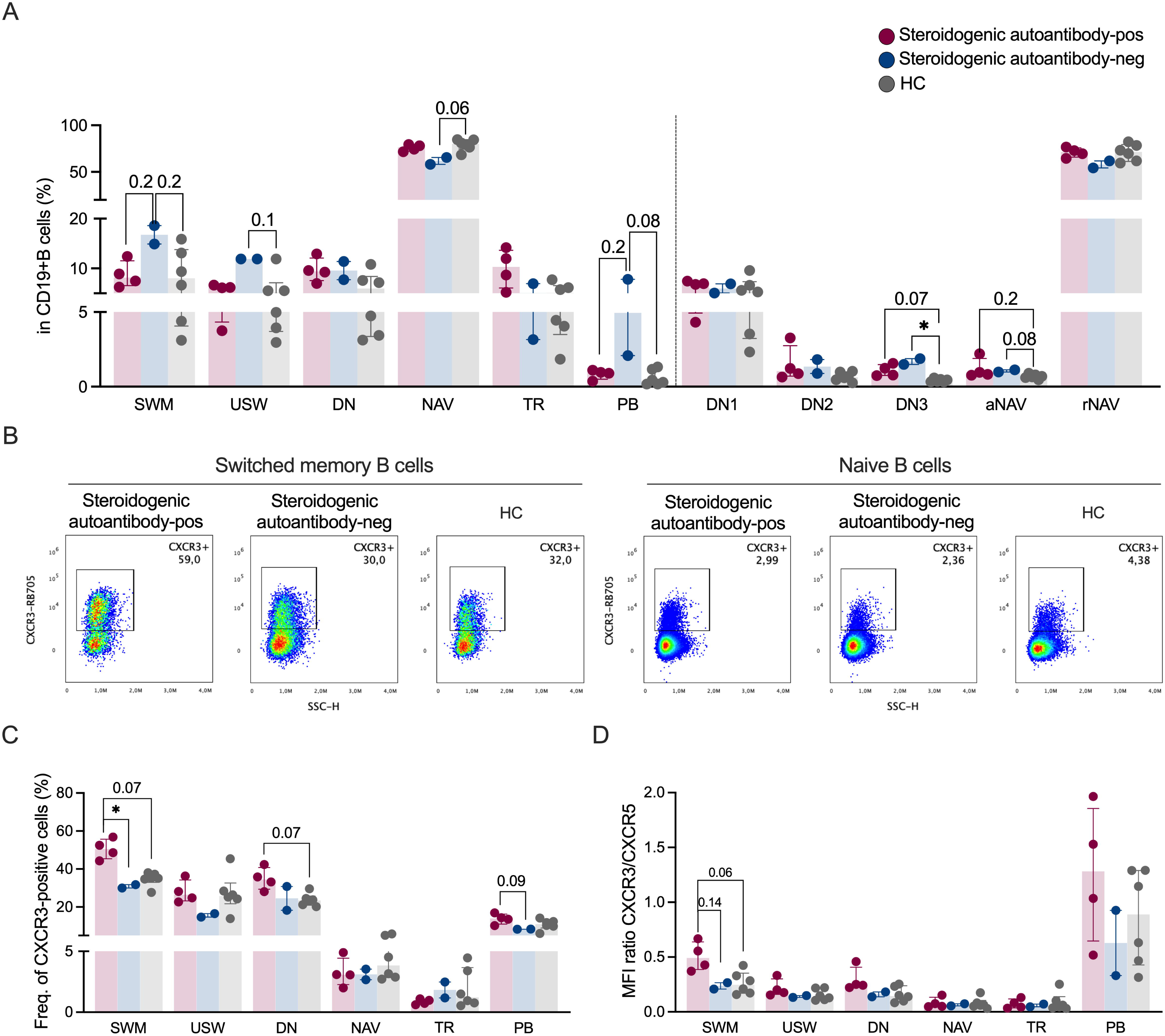
Reduced memory B-cell frequencies with increased CXCR3 expression in patients positive for steroidogenic autoantibodies. (A) Frequencies of SWM, USW, DN, NAV, TR and PB as well as DN (DN1, DN2 and DN3) and NAV (aNAV and rNAV) subsets in patients with steroidogenic autoantibody-positive (n = 4) and-negative (n = 2) and HC (n = 6). (B) Flow cytometric plots representing CXCR3 expression in SWM (left panel) and NAV (right panel) B cells from patient with different subgroups and HC. (C) Frequency of CXCR3-positive cells and (D) MFI ratio of CXCR3 relative to CXCR5 in different B cell subsets across different groups. Red and blue dots indicate patients positive-and negative-for steroidogenic (21OH, 17αOH and/or SCC) autoantibodies, respectively. Data (A, C and D) are analysed by Kruskal-Wallis test and *p* values are corrected by Dunn’s test for multiple comparisons. \**p*< 0.05; \*\**p*< 0.01; \*\*\**p*< 0.001; \*\*\*\**p*< 0.0001.

### CXCR3 upregulation in memory B cells and plasmablasts in steroidogenic autoantibody-positive autoimmune POI patients

Given the reduced frequencies of SWM and PB in steroidogenic autoantibody-positive patients and reported expression of enzymes 17αOH and SCC in ovarian tissues (29, 30), we next investigated whether these subsets displayed tissue-homing phenotype by examining CXCR3 expression (Figure 2B). These patients showed significantly increased CXCR3 expression on SWM compared with other groups (*p* = 0.03). Similar increases were also observed on PB, total DN and DN2 subsets (Figure 2C and S4A). In addition, the CXCR3/CXCR5 ratio was elevated in SWM and DN2 B cells from patients positive for steroidogenic autoantibodies, supporting enhanced tissue-homing capacity of these B cell subsets (Figure 2D, S4B and S4C).

### Alteration of circulating T cell subsets in autoimmune POI

To understand if this enhanced tissue-homing capacity was facilitated by T cells, we assessed the distribution of circulating T cell subsets in all autoimmune POI patients (Figure 3A). When comparing, the frequency of Th1 cells was significantly higher in autoimmune POI compared to HC (*p* = 0.04) (Figure 3B). No differences were found for total CD3+T cells and their subsets, including CD4+, CD8+, γδ+, naïve and memory T cells (Figure 3B and S5).

**Figure 3.**
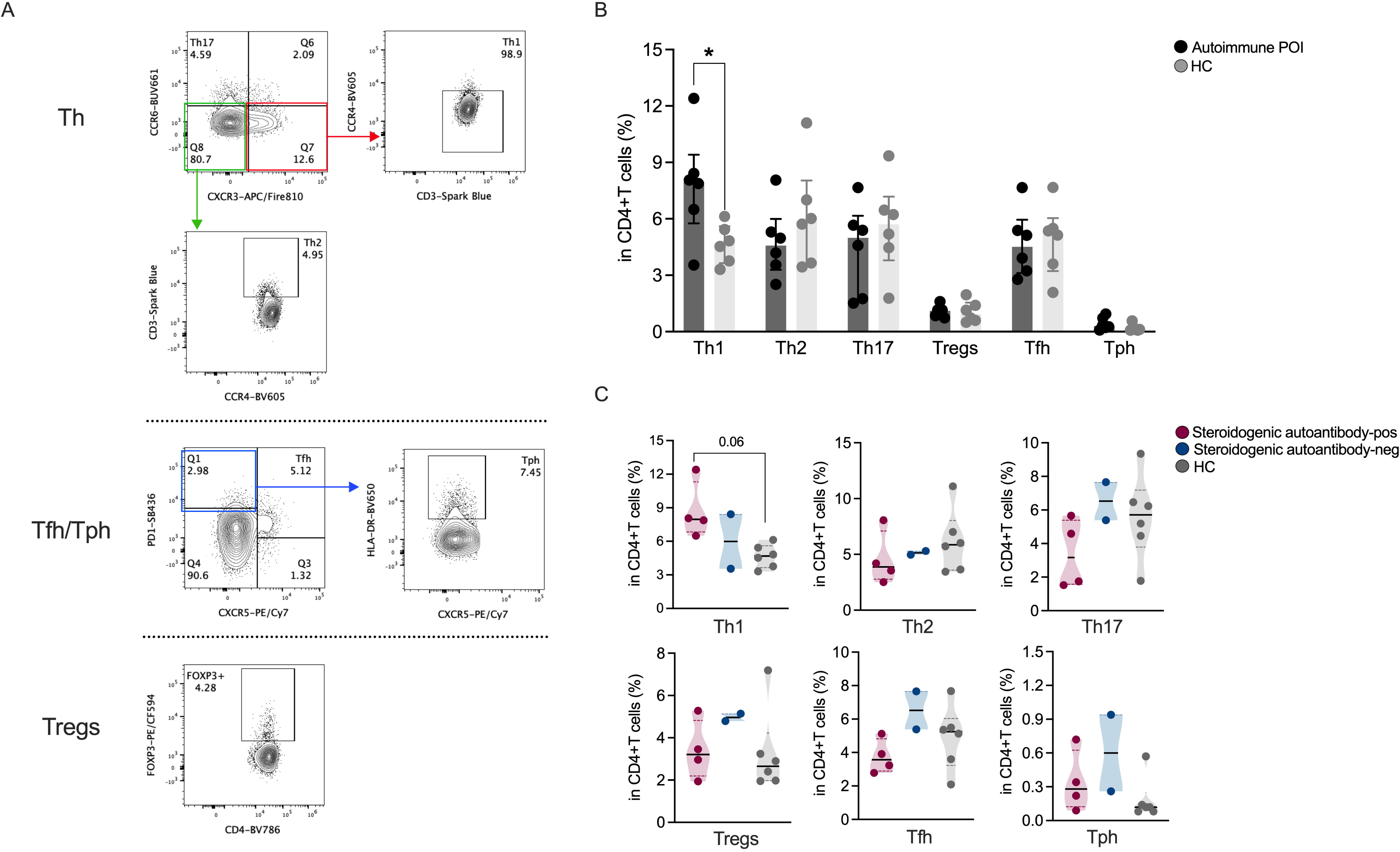
T-cell phenotyping of autoimmune POI patients. (A) Flow cytometric plots showing circulating T helper subsets (Th1: CD3+CD4+CD8-CCR6-CXCR3+CCR4-, Th2: CD3+CD4+CD8-CCR6-CXCR3-CCR4+ and Th17: CD3+CD4+CD8-CCR6+CXCR3-), follicular helper (Tfh: CD3+CD4+CD8-PD1+CXCR5+), peripheral helper-like (Tph-like: CD3+CD4+CD8-CXCR5-PD-1hiHLA-DR+) and regulatory T cells (Tregs: CD3+CD4+CD8-FOXP3+) in autoimmune POI patient. (B) Frequencies of Th1, Th2, Th17, Tregs, Tfh and Tph-like cells in patients (n = 6) and HC (n = 6) as well as (C) in steroidogenic autoantibody-positive and-negative patient subgroups. Red and blue dots indicate patients positive-and negative-for steroidogenic autoantibodies, respectively. Data in B are analysed by Mann-Whitney U test. Data in C are analysed by Kruskal-Wallis test and *p* values are corrected by Dunn’s test for multiple comparisons. \**p*< 0.05; \*\**p*< 0.01; \*\*\**p*< 0.001; \*\*\*\**p*< 0.0001.

We next explored whether the T cell subset distributions differed according to autoantibody status. Despite the absence of significancy, higher frequencies of Th1 cells were found in autoimmune POI setting which seems to increase upon presence of steroidogenic autoantibodies (Figure 3C). This patient group also tended to display higher frequencies of CD3+T cells together with lower frequencies of Th17 and Tfh cells even though significancy was not reached (Figure 3C and S6).

### Association between immune cell subsets and autoantibody levels

To investigate the association between immune cell alterations and autoantibody levels, we performed correlation analyses between frequencies of lymphocyte subsets and levels of anti-21OH, anti-17αOH and anti-SCC antibodies. Correlation heatmap revealed a distinct pattern of associations between B-and T-cell subsets and autoantibody levels (Figure 4A). While SWM frequencies showed only weak positive correlations with autoantibody levels, CXCR3 expression on SWM B cells was more strongly correlated with anti-17αOH (R = 0.9) and anti-SCC (R = 0.71) autoantibody levels, reaching statistical significance only for anti-17αOH (*p* = 0.03) (Figure 4B and 4C). No significant associations were observed between other immune cell subsets and autoantibody levels.

**Figure 4.**
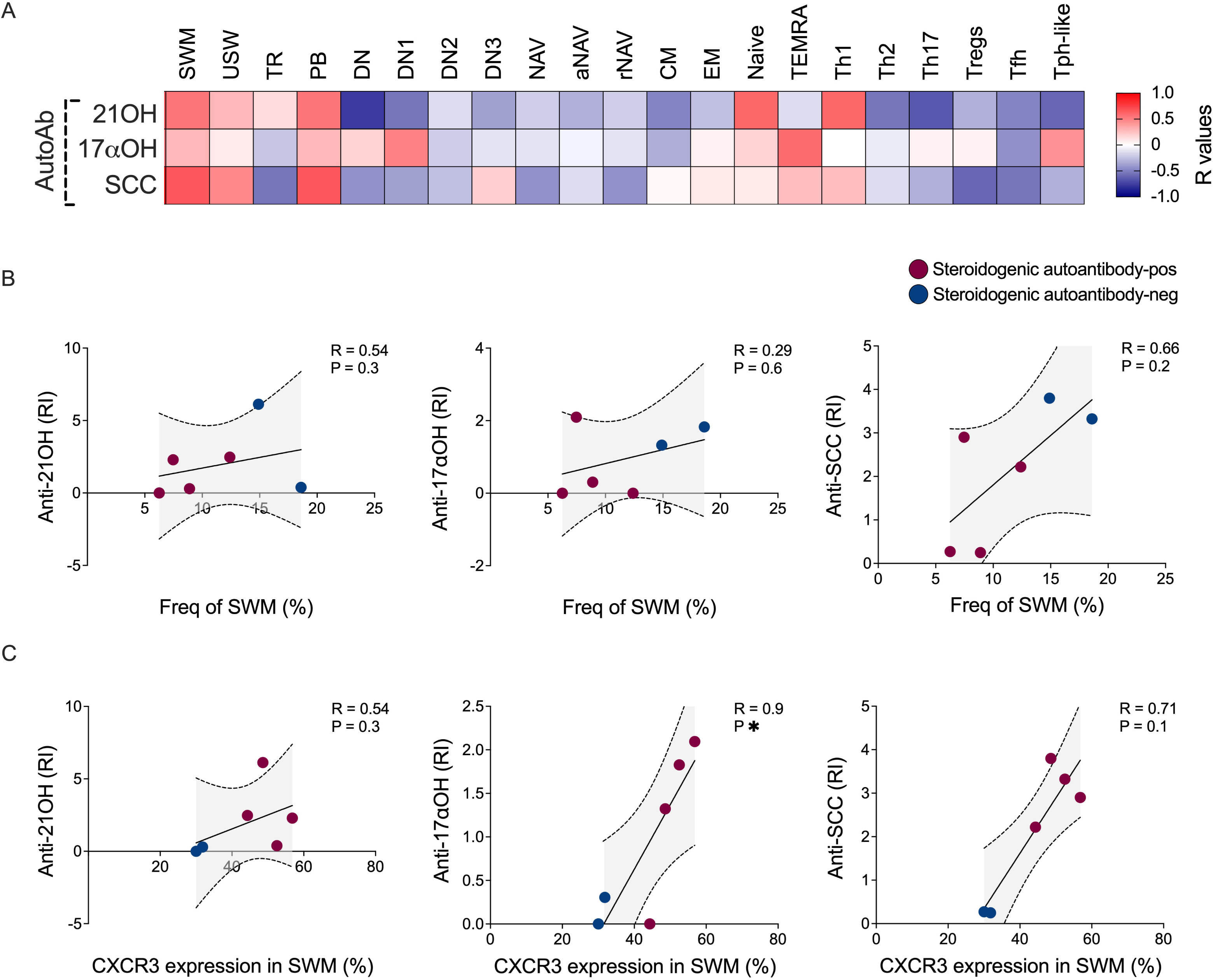
Correlation analysis between different immune cell compartments and autoantibody levels. (A) Correlation matrix heatmap showing the frequencies of B-and T-cell subsets and autoantibody levels (RI: relative index). The strength of the correlation is represented by varying intensity levels (-1 to 1). Correlation analysis between the frequencies of (B) SWM and (C) CXCR3-expressing SWM B cells with anti-21OH, anti-17αOH and anti-SCC antibodies in autoimmune POI patients. Red and blue dots indicate patients positive-and negative-for steroidogenic autoantibodies, respectively. Shaded bands indicate the 95% confidence interval. Correlation analysis was performed using Spearman’s rank coefficient (R) \**p*< 0.05; \*\**p*< 0.01; \*\*\**p*< 0.001; \*\*\*\**p*< 0.0001.

### Changes of B cell subsets after RTX treatment

Patients received RTX following baseline sampling and were monitored longitudinally. Circulating CD19+B cells decreased rapidly after treatment and were significantly reduced compared to baseline. B cell repopulation was observed at 8 months with more substantial repopulation observed at 12 months (Figure 5A). Within the CD19+B cell compartment, we observed a decline in the proportion of memory including SWM, USW and DN (DN1 and DN2 subsets), while NAV and TR B cells increased at 12 months, with enrichment of the elevated NAV B cell compartment accounted from rNAV B cells (Figure 5B and S7A). Intriguingly, a slight increase in PB frequencies was observed at 12 months following RTX (Figure 5B).

**Figure 5.**
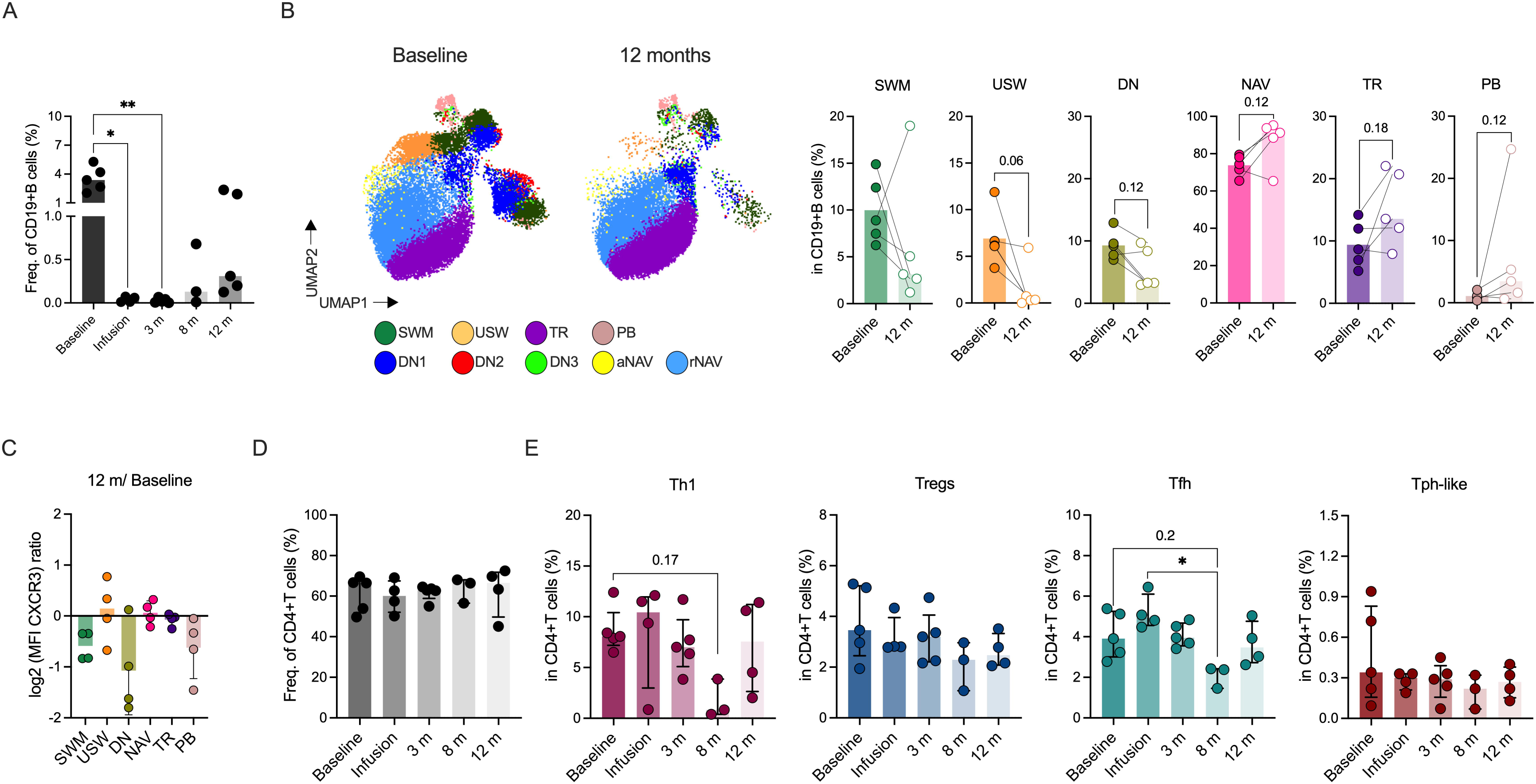
B-and T-cell subsets after rituximab treatment. (A) Frequency of CD19+B cells at baseline and follow-up time points (second infusion, 3, 8 and 12 months) in autoimmune POI patients (n = 5). (B) UMAP of CD19+B cells showing different B cell subsets at baseline and 12 months following rituximab (RTX) treatment (left panel) and frequencies of SWM, USW, DN, NAV, TR and PB at baseline and 12 months post RTX (right panel). (C) Bar graph showing fold changes in the MFI of CXCR3 expression on different B cell subsets at 12 months after treatment relative to baseline. Frequencies of (D) CD4+T cells and (E) their subsets (Th1, Tregs, Tfh and Tph-like) at different time points. Data are represented as median and the comparison of baseline and follow up samples are analysed by the Wilcoxon-matched pairs signed rank test and multiple comparisons are analysed by Kruskal-Wallis test and *p* values are corrected by Dunn’s test. \**p*< 0.05; \*\**p*< 0.01; \*\*\**p*< 0.001; \*\*\*\**p*< 0.0001.

We next assessed CXCR3 expression across different B cell compartments after RTX treatment in comparison to baseline (Figure 5C). SWM, DN and PB subsets showed decreased CXCR3 expression at 12 months post RTX, indicating that these reconstituting B cells may have reduced migratory potential.

### T cell subset frequencies after RTX treatment

As T cells play a role in supporting B cell responses, we further explored longitudinal changes in CD4+T cell subsets at baseline, second infusion, 3, 8 and 12 months following RTX treatment. Overall CD4+T cells remained stable across all time points (Figure 5D and S8). Analysis of CD4+T cell subsets revealed a trend toward reduced frequencies of Th1 and Tfh cells after RTX treatment, with the lowest frequencies observed at 8 months, although statistical significance was reached only for Tfh cells (*p* = 0.03) (Figure 5E). These cell subsets subsequently increased and approached baseline levels at 12 months. Frequencies of Tregs, Tph-like, Th2 and Th17 cells were comparable in all time-points (Figure 5E and S7B).

Among all patients, three were responders after RTX treatment. Their patterns of B-and T-cell dynamics were comparable to those observed in the overall cohort (Figure S9 and S10).

## Discussion

In this pilot study, we performed an in-depth immunophenotyping of circulating B cell and T cell subsets in patients with autoimmune POI undergoing B-cell depleting RTX therapy in conjunction with ovarian stimulation. As recently published, a subset of patients underwent successful pregnancies (13), suggesting that a transient reset/retune of the underlying autoimmunity allowed a time window for ovarian function to recover. When assessing the circulating lymphocytes before RTX we could not detect any gross disturbances among the classical B and T cell subsets. However, we did observe elevated levels of the tissue homing chemokine receptor CXCR3 on switched memory (SWM) B cells compared with healthy controls, especially in the patients positive for steroidogenic autoantibodies (21OH, 17αOH and/or SCC), targeting key steroidogenic enzymes, including those expressed in the ovaries. Moreover, such CXCR3 expression positively associated with anti-17αOH and anti-SCC levels, suggesting a potential tissue-homing capacity. In parallel, Th1 cells (as defined by CXCR3 expression) were simultaneously expanded in the autoimmune POI patients, with a similar trend observed in the steroidogenic autoantibody-positive group. Collectively, the findings support a concept where POI patients with autoimmunity targeting the ovaries have antigen-experienced B cells that migrate to affected ovarian tissues and ensuing inflammation hamper their physiological function.

POI patients are a heterogeneous group with varying underlying causes including genetics, infections as well as different autoimmune diseases (2, 3). Our pilot data and the recent proof-of-concept study (13) collectively suggest that POI patients with autoimmunity directly involving the ovaries, i.e. autoantibodies to steroidogenic enzymes could benefit from therapeutic targeting of the adaptive immune system. Both 17αOH and SCC are expressed by steroidogenic cells in the ovaries (29, 30). 21OH is classically regarded as an adrenal-specific enzyme. However, 21OH-derived steroids have been detected in human luteinized granulosa cells (31).

We report an increased proportion of CXCR3+B cells in the circulation of patients prior to RTX treatment. CXCR3 is a tissue homing chemokine receptor which can be upregulated e.g. following exposure to the Th1-associated cytokine IFN-γ (25,26). Indeed, we also observed an increased proportion of CXCR3+T cells which are commonly used as a proxy for Th1 cells. As a key mediator of lymphocyte trafficking, CXCR3 mediates migration in response to the inflammatory chemokines CXCL9, CXCL10 and CXCL11 (32, 33), whose circulating levels have been reported to be elevated in POI (34) and autoimmune Addison’s disease (35,36). Histological studies of autoimmune oophoritis have shown lymphocytic infiltrates, including predominantly T cells together with B cells and plasma cells in affected tissue (37). Although comparable histological data are lacking from the autoimmune POI patients in our study, our findings support the concept that B cells recognising ovarian antigens migrate to inflamed ovarian tissue.

In our longitudinal setup, we could validate B cell repopulation at 8-12 months, and an expected reset with primarily naïve B cells and hence a reduction of CXCR3 expressing, tissue homing B cells. In contrast, total CD4+T cells and their subsets (which are not directly affected by RTX) remained initially stable, with a delayed transient decline of Th1 and Tfh cells. Together, these findings may indicate a downstream effect of helper T cell responses dependent on B cell interaction, although we cannot formally exclude an influence from the ovarian stimulation performed between months 4-6 which we could not control for.

Compared to other autoantibody positive diseases treated with RTX, the overall B cell aberrations observed in this patient group were modest. This possibly reflects that these patients have more local rather than systemic autoimmunity. A trend toward lower expression of immune checkpoint molecule BTLA on B cells in autoimmune POI, in line with reports from other autoimmune diseases (23,38), may reflect altered immune regulation. Still, we found an enrichment of aNAV and DN3 B cell subsets, with a similar trend for DN2 cells, changes which are considered markers of ongoing autoreactive immune responses in SLE, RA and Sjögren’s disease (21,23,24,39,40). These are conditions where next-generation anti-CD20 antibodies (e.g., obinutuzumab), bispecific antibodies and chimeric antigen receptor (CAR) T cell approaches are currently being explored (41–43) both to achieve deeper B cell depletion including in tissues and to minimize the appearance of anti-drug antibodies which may lead to therapeutic loss (44,45). Herein, patients with autoimmune POI received a single course of RTX consisting of two infusions 14 days apart. Whether more sustained B cell depletion would provide additional clinical benefit remains to be determined.

The main limitation of this study is the small number of patients available for analysis, reflecting the rarity of autoimmune POI. Additionally, a double-blind randomized clinical trial is needed, and planned, to validate the observed clinical outcome. From an immunological perspective, it would also be of considerable interest to access tissue or follicular fluid to assess the extent to which ovaries are infiltrated by the adaptive immune system. We also did not have the opportunity to study antigen-specific autoreactive B cells to validate whether they are enriched for CXCR3 expression. However, we do believe that our findings contribute to a better understanding of the underlying pathophysiology of the disease and provide a foundation for future studies in larger cohorts.

In conclusion, we present an expanded migratory phenotype of antigen-experienced B cells characterized by upregulation of CXCR3 in autoimmune POI patients with steroidogenic autoantibodies. Together with the positive association with autoantibody levels and expansion of Th1 cells, these findings support the concept that B cells may be recruited to the ovary within a Th1-polarized inflammatory environment. Reduction in CXCR3-expressing B cells after RTX further suggests that depletion may alter immune cell trafficking, providing a potential immunological mechanism underlying the previously observed improvement in ovarian function.

## Acknowledgements

We thank all patients with autoimmune POI for their willingness to participate in this study.

## Author contributions

Patient recruitment and revision of clinical data: ALH SB, SB and IG. Study concept and design: KW, AH, KA, FF, OK, ALH, VM and IG. Acquisition of data: KW, AH and KA. Analysis of data: KW, AH, MAG and KA. Interpretation of data: KW, AH, KA, FF, VM and IG. Manuscript writing: KW, AH, KA, FF, SB, SB, OK, ALH, VM and IG. All authors read and approved the final manuscript.

## Data availability statement

The data generated during and/or analysed during the current study are available from the corresponding author on request.

## Abbreviations

17αOH: 17α-hydroxylase
21OH: 21-hydroxylase
AChR: Acetylcholine receptor
CXCR3: C-X-C motif chemokine receptor 3
CXCR5: C-X-C motif chemokine receptor 5
CM: Central memory
DN: Double negative
EM: Effector memory
HC: Healthy controls
MS: Multiple sclerosis
NAV: Naïve
PB: Plasmablasts
POI: Premature ovarian insufficiency
RA: Rheumatoid arthritis
Treg: Regulatory T cell
RTX: Rituximab
SCC, CYP11A1: Side-chain cleavage enzyme
SWM: Switched memory
SLE: Systemic lupus erythematosus
Tfh: T follicular helper
Tph: T peripheral helper
Th: T helper
TEMRA: Terminally differentiated effector memory
TPO: Thyroid peroxidase
TR: Transitional
USW: Unswitched memory

