## Supplementary file for "Immunophenotyping of B- and T-cell alterations in patients with autoimmune premature ovarian insufficiency following rituximab treatment"

**Supplementary table 1** Flow cytometry panel used for B cell phenotyping

| Antibody | Fluorophore | Clone | Company | Dilution | Catalogue No. |
| --- | --- | --- | --- | --- | --- |
| eFluor506 | LD | 1:1000 |  | Thermofisher | 65-0866-14 |
| eFluor506 | CD3 | 1:25 | UCHT1 | Invitrogen | 69-0038-42 |
| eFluor506 | CD14 | 1:25 | 61D3 | Invitrogen | 69-0149-42 |
| BV480 | IgD | 1:200 | IA6-2 | BD | 566138 |
| BUV395 | CD20 | 1:50 | 2H7 | BD | 563782 |
| APC/Fire810 | CD27 | 1:100 | QA17A18 | Biolegend | 393214 |
| BUV563 | CD38 | 1:400 | HB7 | BD | 741446 |
| BUV805 | CD21 | 1:400 | B-ly4 | BD | 742008 |
| BUV496 | CD24 | 1:25 | ML5 | BD | 741143 |
| SB436 | IgM | 1:50 | SA-DA4 | Invitrogen | 62-9998-42 |
| VioBlue | IgA | 1:400 | IS11-8E10 | Miltenyi | 130-113-479 |
| BV421 | IgG | 1:50 | G18-145 | BD | 562581 |
| BV570 | CD19 | 1:25 | HIB19 | Biolegend | 302236 |
| BUV615 | CD45RB | 1:800 | MT4 (6B6) | BD | 751482 |
| BV711 | CD138 | 1:25 | MI15 | Biolegend | 356522 |
| BV750 | CXCR5 | 1:50 | J24D4 | Biolegend | 356942 |
| BV785 | CD73 | 1:100 | AD2 | Biolegend | 344028 |
| PE/Cy5 | CD95 | 1:800 | DX2 | Biolegend | 305610 |
| BV650 | HLA-DR | 1:400 | L243 | Biolegend | 307650 |
| APC/Fire750 | CD11c | 1:400 | S-HCL-3 | Biolegend | 371509 |
| PE/Fire810 | CCR7 | 1:100 | G043H7 | Biolegend | 353269 |
| RB545 | CD72 | 1:200 | J4-117 | BD | 756280 |
| AF647 | BTLA | 1:50 | MIH26 | Biolegend | 344519 |
| PE | CD360 (IL-21R) | 1:50 | 17A12 | Biolegend | 359506 |
| BUV737 | SLAMF7 | 1:25 | 235614 | BD | 750833 |
| BB515 | CD86 | 1:50 | 2331 (FUN-1) | BD | 564544 |
| BV605 | CD1c | 1:200 | L161 | Biolegend | 331537 |
| RB744 | CD71 | 1:100 | M-A712 | BD | 757854 |
| APC | BCMA | 1:25 | 19F2 | Biolegend | 357506 |
| RB705 | CXCR3 | 1:100 | 1C6 | BD | 570554 |

**Supplementary table 2** Flow cytometry panel used for T cell phenotyping

| **Antibody** | **Fluorophore** | **Clone** | **Company** | **Dilution** | **Catalogue No.** |
| --- | --- | --- | --- | --- | --- |
| APC/FIRE810 | CXCR3 | 1:25 | G025H7 | Biolegend | 353762 |
| BUV661 | CCR6 | 1:25 | 11A9 | BD | 750696 |
| eFluor506 | CD14 | 1:25 | 61D3 | ThermoFisher | 69-0149-42 |
| eFluor506 | CD19 | 1:25 | HIB19 | ThermoFisher | 69-0199-42 |
| Spark blue 550 | CD3 | 1:50 | SK7 | Biolegend | 344852 |
| SB436 | PD-1 | 1:50 | eBioJ105 | ThermoFisher | 62-2799-42 |
| PE/Cy7 | CXCR5 | 1:50 | J252D4 | Biolegend | 356923 |
| PE | ICOS | 1:50 | DX29 | BD | 557802 |
| Pacific blue | CD127 | 1:50 | A019D5 | Biolegend | 351306 |
| BUV496 | CD27 | 1:50 | O323 | BD | 751678 |
| BV605 | CCR4 | 1:50 | 1G1 | BD | 562906 |
| PE/FIRE810 | CCR7 | 1:50 | G043H7 | Biolegend | 353269 |
| BV786 | CD4 | 1:100 | SK3 | BD | 563877 |
| BUV395 | GPR56 | 1:100 | CG4.rMAb | BD | 752707 |
| BV480 | CD25 | 1:100 | BC96 | BD | 567488 |
| AF700 | CD45Ra | 1:100 | HI100 | Biolegend | 304120 |
| BV711 | TCR$\gamma\delta$ | 1:100 | 11F2 | BD | 568490 |
| BV750 | CD69 | 1:100 | FN50 | BD | 747522 |
| PE/Cy5 | CD137 | 1:100 | 4B41 | Biolegend | 309808 |
| BUV737 | CD40L | 1:100 | TRAP1 | BD | 748983 |
| RB780 | TIGIT | 1:200 | TgMab-2 | BD | 569940 |
| RB705 | CCR2 | 1:200 | LS132.1D9 | BD | 757633 |
| APC/FIRE750 | CD8 a +b | 1:400 | SK1 | Biolegend | 344745 |
| BUV563 | CD38 | 1:400 | HB7 | BD | 741446 |
| BV650 | HLA-DR | 1:400 | L243 | Biolegend | 307650 |
| BUV805 | CTLA-4 | 1:100 | BNI3 | BD | 569654 |
| AF647 | GZMK | 1:50 | GM26E7 | Biolegend | 370503 |
| FITC | GZMB | 1:50 | QA18A28 | Biolegend | 372205 |
| PE/Dazzle594 | FOXP3 | 1:33 | 206D | Biolegend | 320125 |

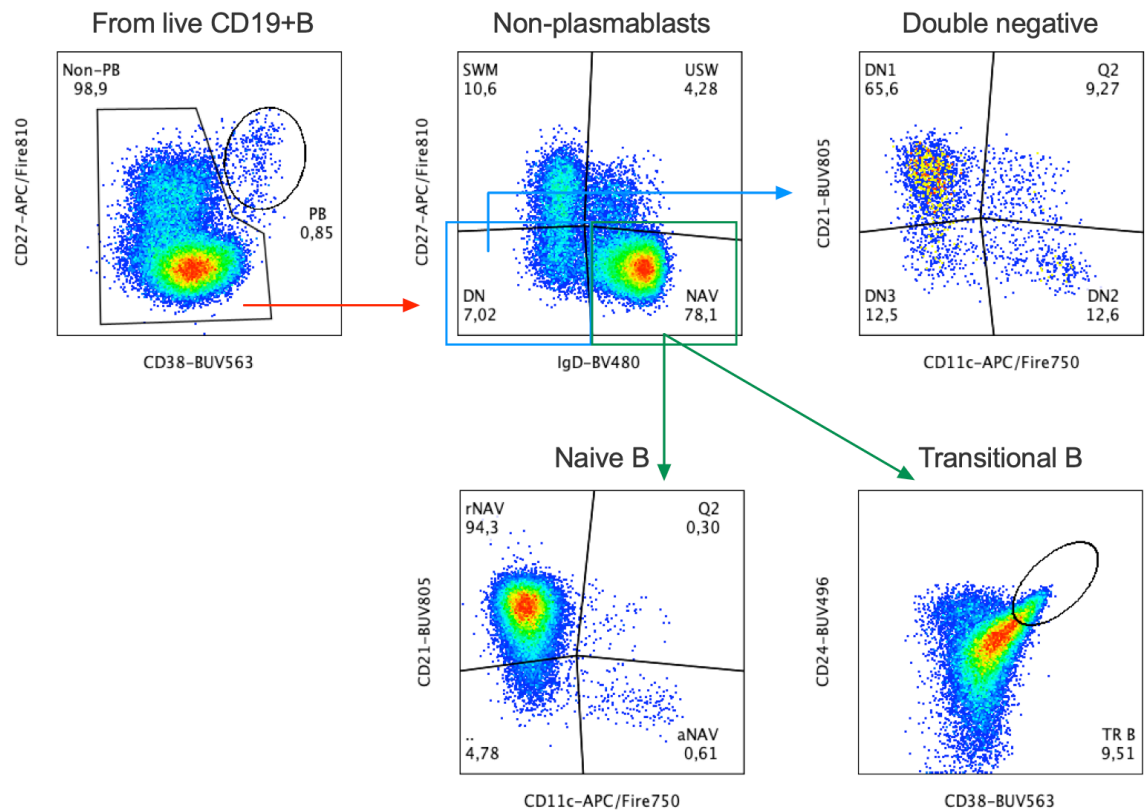

**Supplementary figure 1** Phenotype of circulating B cells in autoimmune POI patients. Representative gating strategies of CD19+B-cells from one patient. The five independent subpopulations were identified: plasmablast (PB: CD19+CD27<sup>hi</sup>CD38<sup>hi</sup>), switched memory (SWM: CD19+CD27+IgD<sup>-</sup>), unswitched memory (USW: CD19+CD27+IgD<sup>+</sup>), transitional (TR: CD19+CD27-IgD+CD24<sup>hi</sup>CD38<sup>hi</sup>), double negative (DN: CD19+CD27-IgD<sup>-</sup>) and naïve B cells (NAV: CD19+CD27-IgD<sup>+</sup>). The subsets of DN and NAV B-cells were classified and grouped as double negative 1 (DN1: CD19+CD27-IgD-CD21+CD11c<sup>-</sup>), double negative 2 (DN2: CD19+CD27-IgD-CD21-CD11c<sup>+</sup>), double negative 3 (DN3: CD19+CD27-IgD-CD21-CD11c<sup>-</sup>), resting naïve (rNAV: CD19+CD27-IgD+CD21+CD11c<sup>-</sup>) and activated naïve (aNAV: CD19+CD27-IgD+CD21-CD11c<sup>+</sup>).

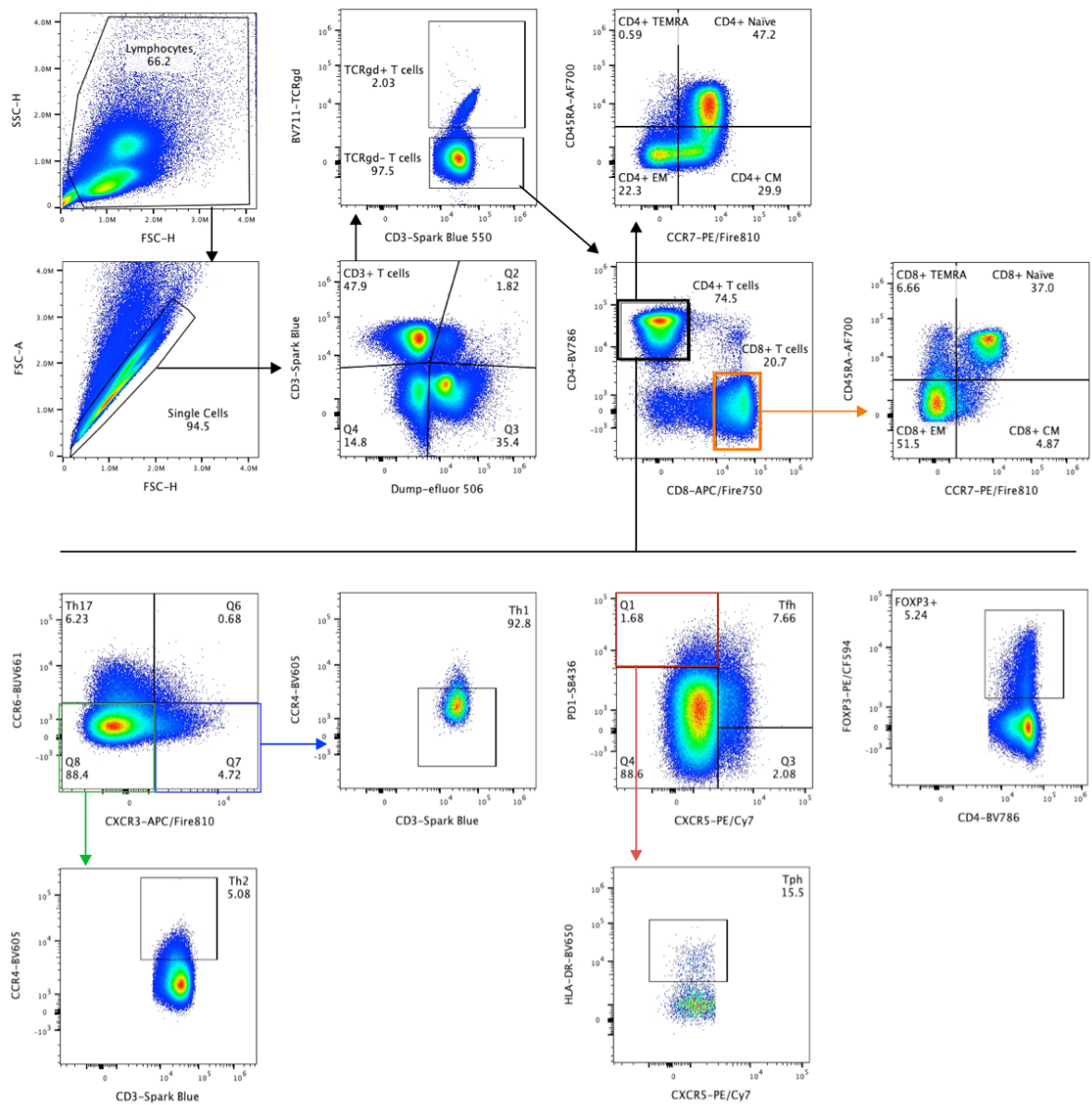

**Supplementary figure 2** Gating strategy of circulating T cell in a representative autoimmune POI patient. CD3<sup>+</sup> T cells were subdivided into TCRγδ<sup>+</sup> and TCRγδ<sup>-</sup> T cells. CD4<sup>+</sup> and CD8<sup>+</sup> T cells were identified from the TCRγδ<sup>-</sup> T cell pool together with their subsets. CD4<sup>+</sup> subsets: T helper cell 1 (Th1: CCR6-CXCR3+CCR4-), Th2 (CCR6-CXCR3-CCR4+), Th17 (CCR6+CXCR3-), T peripheral helper cell (Tph: PD1hiCXCR3-HLA-DR+), T follicular helper cell (Tfh: PD1+CXCR5+), regulatory T cell (Treg: FOXP3+), central memory (CM: CCR7+CD45RA-), effector memory (EM: CCR7-CD45RA-), terminally differentiated effector memory (TEMRA: CCR7-CD45RA+), and Naïve (NAV: CCR7+CD45RA+). CM, EM, TEMRA, and NAV T cells were also identified within the CD8<sup>+</sup> T cell pool.

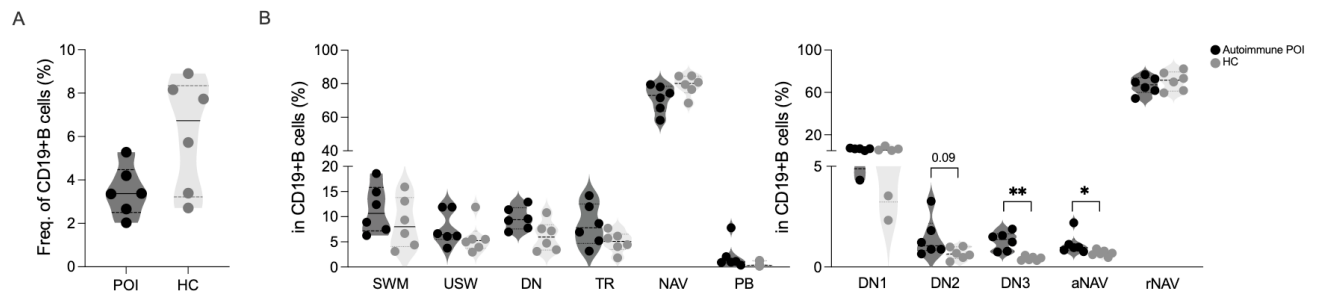

**Supplementary figure 3** Frequencies of (A) CD19+B cells, (B) SWM, USW, DN, TR, NAV and PB (left panel) as well as DN1, DN2, DN3, aNAV and rNAV (right panel) in autoimmune POI patients (n = 6) compared to healthy controls (HC, n = 6). Data are represented as median and are analysed by Mann-Whitney U test. \*$p < 0.05$; \*\*$p < 0.01$; \*\*\*$p < 0.001$; \*\*\*\*$p < 0.0001$.

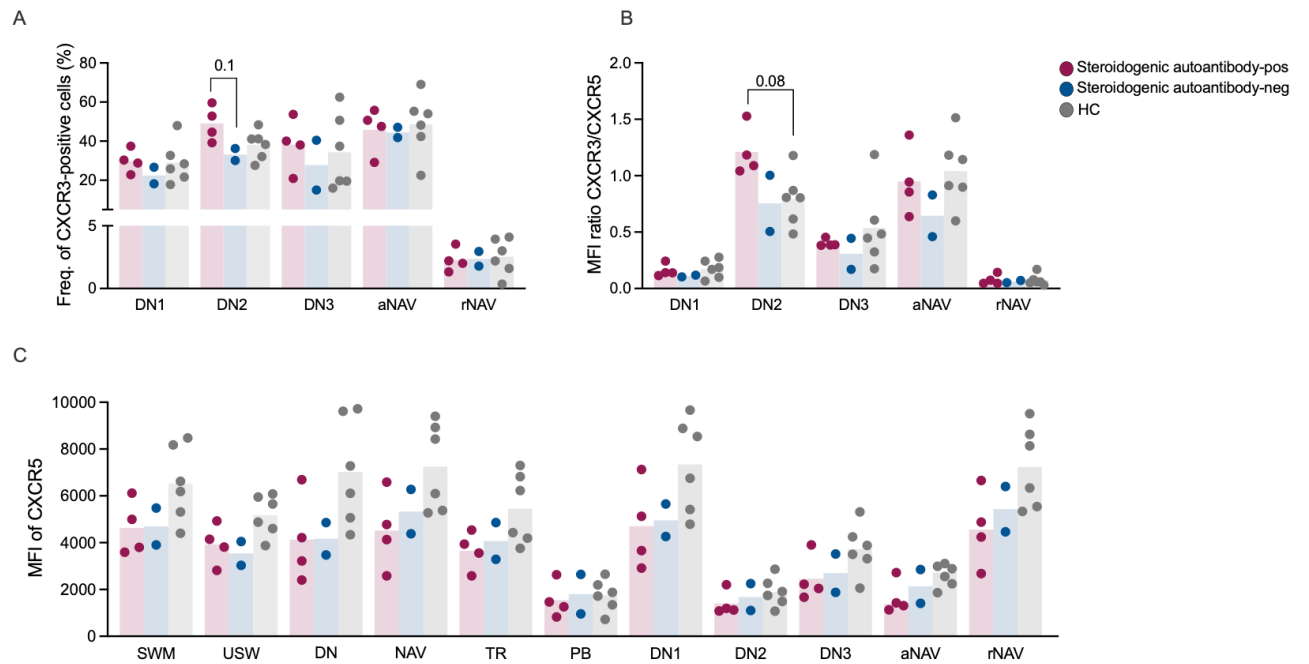

**Supplementary figure 4** (A) Frequency of CXCR3-positive cells and (B) MFI ratio of CXCR3 relative to CXCR5 in DN1, DN2, DN3, aNAV and rNAV B cells in patients with steroidogenic autoantibody-positive and -negative and HC. (C) MFI of CXCR5 in all B cell subsets including SWM, USW, DN, NAV, TR, PB, DN and NAV subsets across different groups. Data are analysed by Kruskal-Wallis test and $p$ values are corrected by Dunn's test for multiple comparisons. \*$p < 0.05$; \*\*$p < 0.01$; \*\*\*$p < 0.001$; \*\*\*\*$p < 0.0001$.

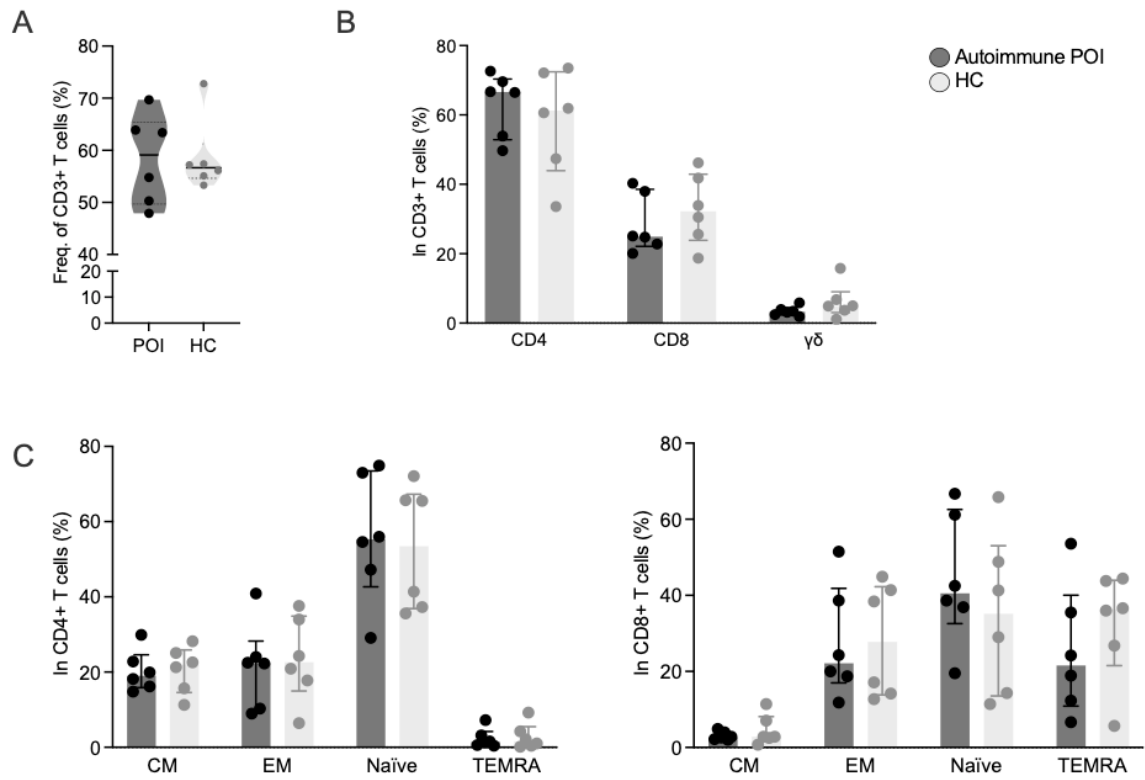

**Supplementary figure 5** Frequencies of (A) CD3+T cells, (B) CD4, CD8, and $\gamma\delta$ T cells as well as both (C) CD4+ and CD8+CM, EM, NAV T cells in autoimmune POI patients (n=6) compared to HC (n=6). Data are represented as median and analysed by Mann-Whitney U test. \*$p < 0.05$; \*\*$p < 0.01$; \*\*\*$p < 0.001$; \*\*\*\*$p < 0.0001$.

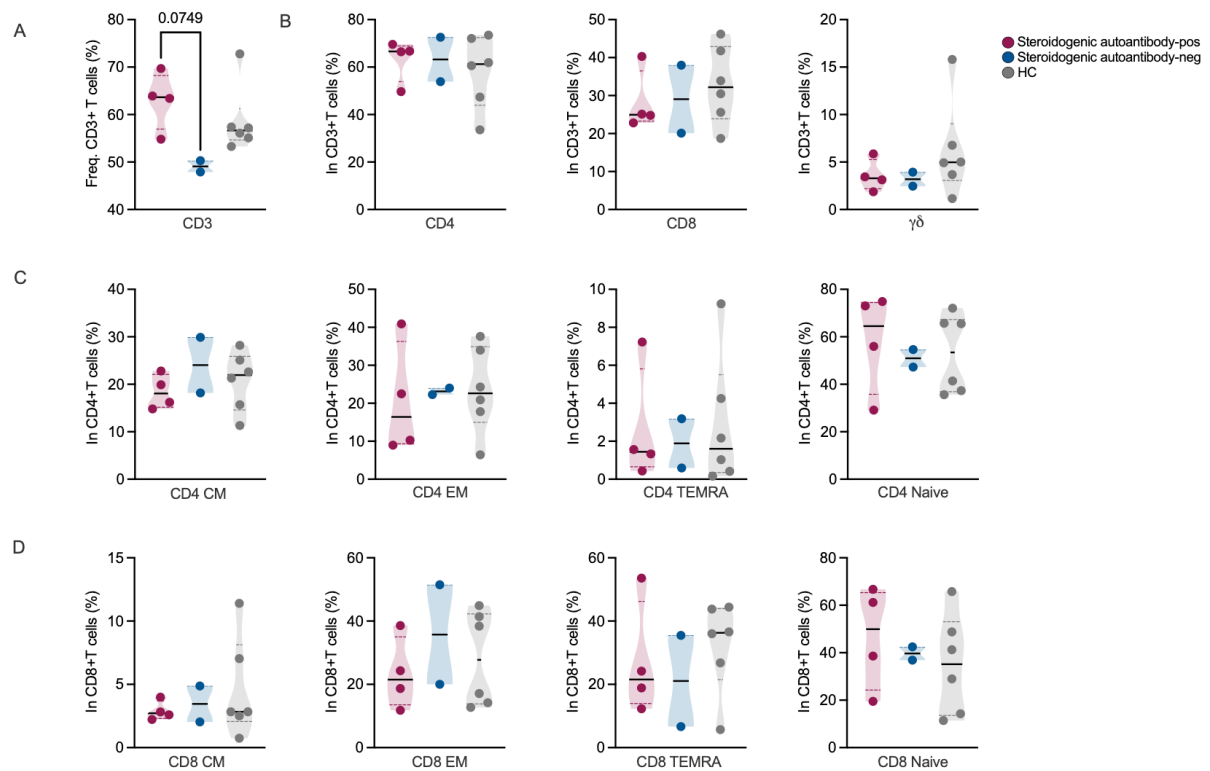

**Supplementary figure 6** Frequencies of (A) CD3+T cells, (B) CD4+, CD8+, and $\gamma\delta$+T cells as well as both (C) CD4+ and (D) CD8+CM, EM, NAV T cells in patients with steroidogenic autoantibodies-positive (n = 4) and -negative (n=2) and HC (n=6). Data are analysed by Kruskal-Wallis test and *p* values are corrected by Dunn's test for multiple comparisons. \**p*< 0.05; \*\**p*< 0.01; \*\*\**p*< 0.001; \*\*\*\**p*< 0.0001.

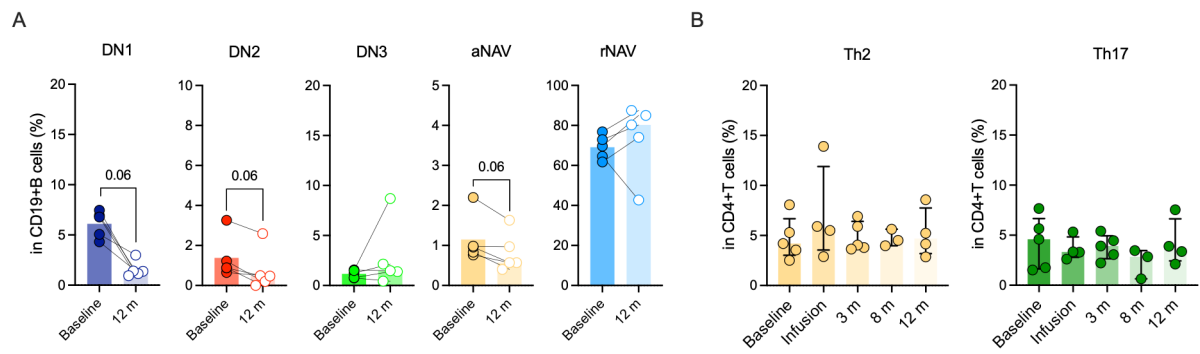

**Supplementary figure 7** (A) Frequencies of DN1, DN2, DN3, aNAV and rNAV B cells at baseline and 12 months post RTX in autoimmune POI patients ($n = 6$). (B) Frequencies of Th2 and Th17 cells in patients at different time points. Data are represented as median and the comparison of baseline and follow up samples are analysed by the Wilcoxon-matched pairs signed rank test and data from multiple timepoints are analysed by Kruskal-Wallis test and $p$ values are corrected by Dunn's test for multiple comparisons. \*$p < 0.05$; \*\*$p < 0.01$; \*\*\*$p < 0.001$; \*\*\*\*$p < 0.0001$.

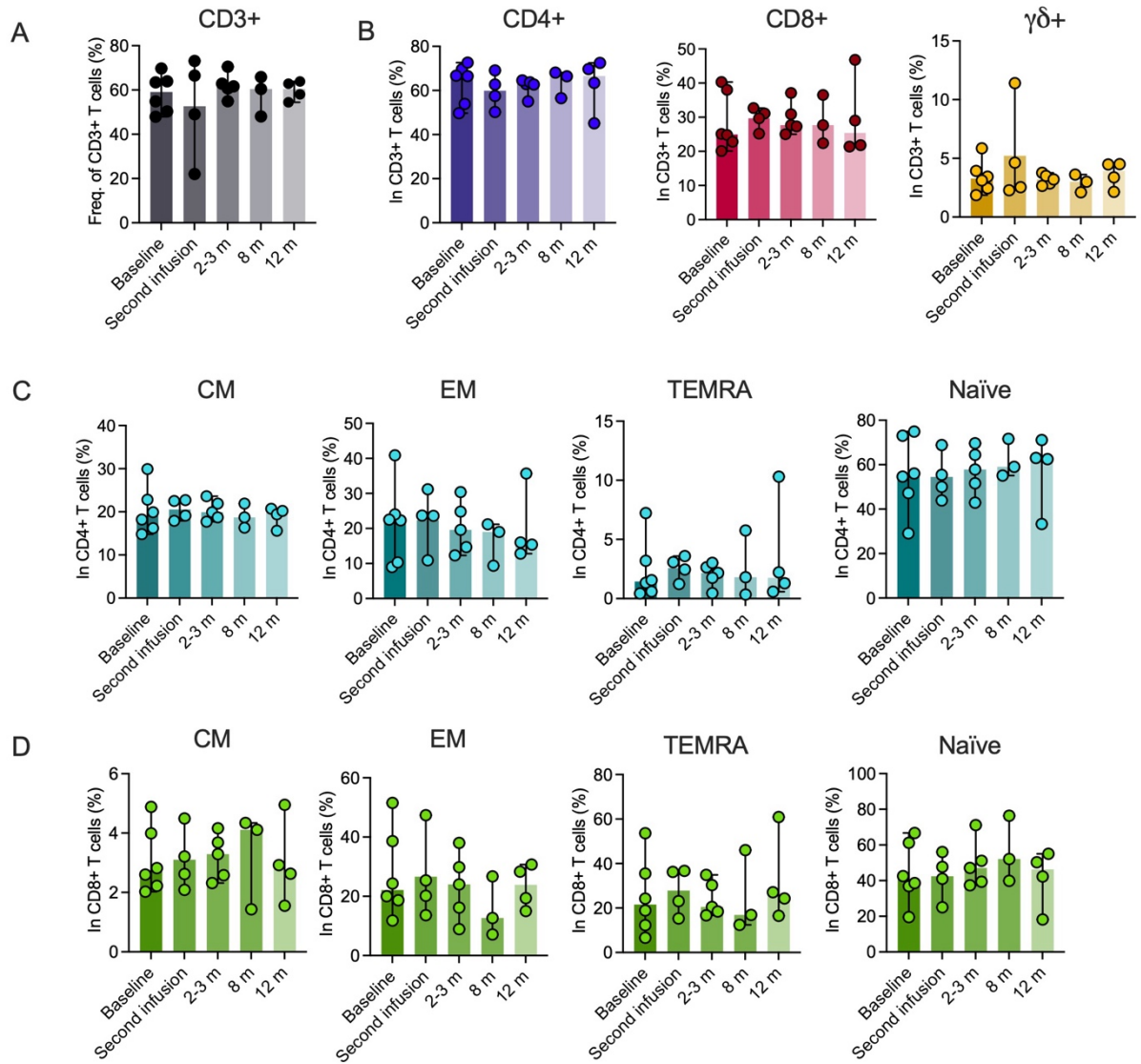

**Supplementary figure 8** Frequencies of (A) CD3+ T cells, (B) CD4, CD8, and $\gamma\delta$ T cells as well as both CD4+ and CD8+CM, EM, NAV T cells at baseline and up till 12 months post RTX treatment in autoimmune POI patients (n = 6). Data are analysed by Kruskal-Wallis test and *p* values are corrected by Dunn's test for multiple comparisons. \**p* < 0.05; \*\**p* < 0.01; \*\*\**p* < 0.001; \*\*\*\**p* < 0.0001.

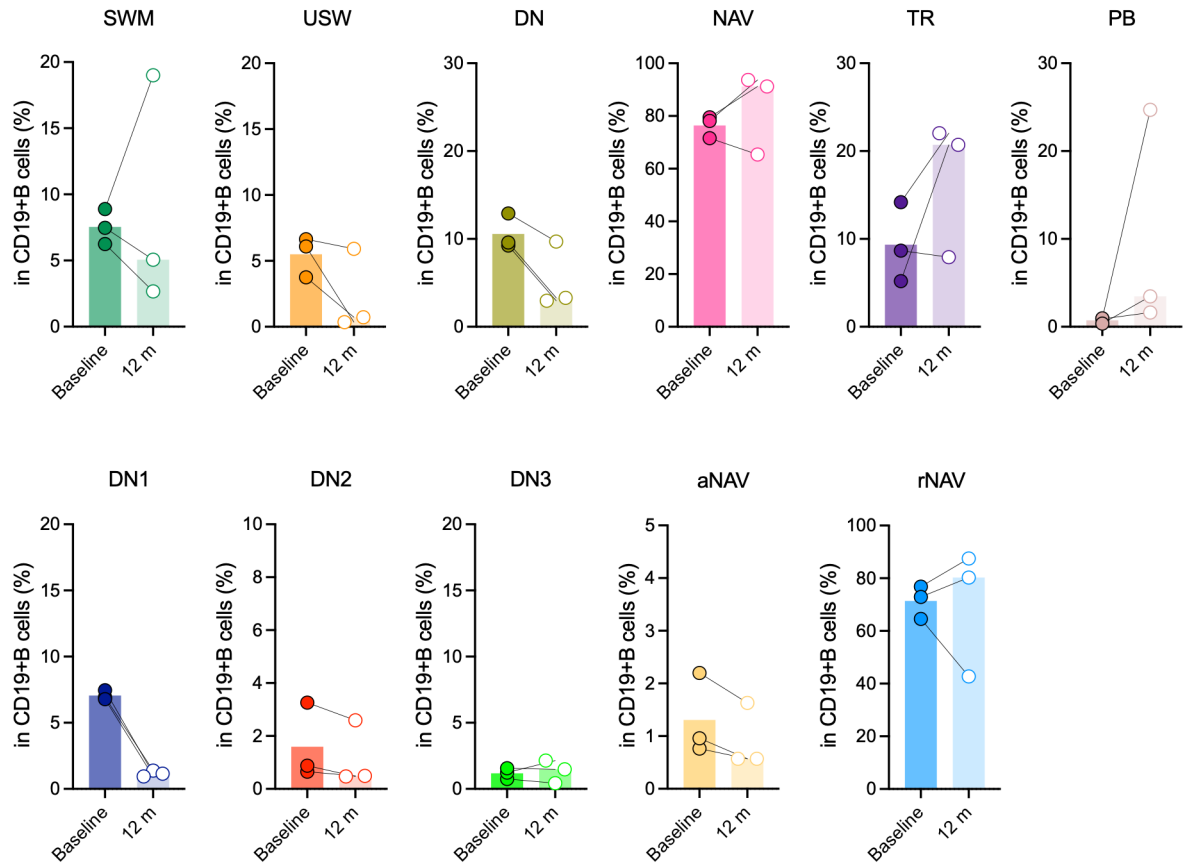

**Supplementary figure 9 (A)** Frequencies of SWM, USW, DN, NAV, TR, PB, DN1, DN2, DN3, aNAV and rNAV B cells at baseline and 12 months post RTX in autoimmune POI patients who had positive fertility outcome (responder, $n = 3$). Data are represented as median and the comparison of baseline and follow up samples are analysed by the Wilcoxon-matched pairs signed rank test. \*$p < 0.05$; \*\*$p < 0.01$; \*\*\*$p < 0.001$; \*\*\*\*$p < 0.0001$.

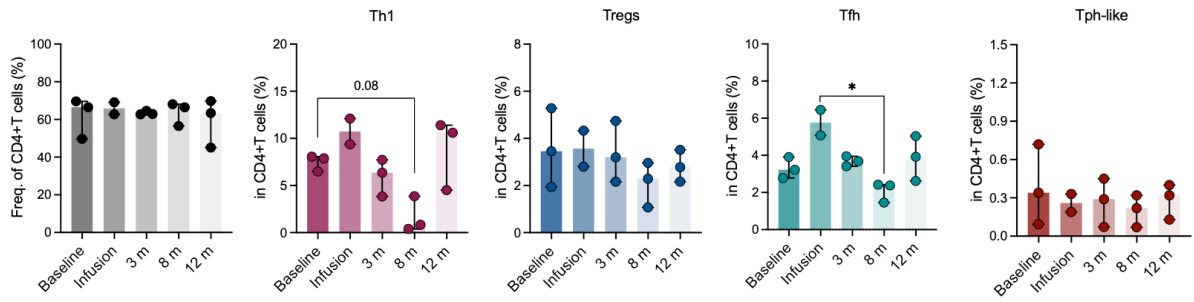

**Supplementary figure 10** Frequencies of overall CD4+T cells and their subsets (Th1, Tregs, Tfh and Tph-like) at baseline and 12 months post RTX in autoimmune POI patients who had positive fertility outcome (responder, n = 3). Data are represented as median and the comparison of baseline and follow up samples are analysed by the Wilcoxon-matched pairs signed rank test. \*$p < 0.05$; \*\*$p < 0.01$; \*\*\*$p < 0.001$; \*\*\*\*$p < 0.0001$.
